# Long-term risk of repeat hospital admission involving self-harm by Aboriginal and non-Aboriginal people: A retrospective cohort study

**DOI:** 10.1101/2023.08.08.23293862

**Authors:** Bernard Leckning, Rohan Borschmann, Tanja Hirvonen, Sven R. Silburn, Steven Guthridge, Gary W. Robinson

## Abstract

**Background:** Identify risk factors for repeat hospitalisation involving self-harm by Aboriginal and non-Aboriginal people in the Northern Territory (NT), Australia.

**Methods:** A retrospective cohort study of hospitalisations involving suicidal ideation and/or self-harm between 1 July 2001 and 31 December 2013 followed up to 31 December 2018. Survival analyses identified demographic and clinical characteristics associated with repeat hospitalisation involving self-harm.

**Results:** The risk of repeat hospitalisation involving self-harm was higher (HR 1.39; 95% CI: 1.22-1.59) amongst Aboriginal (n=2,304) than non-Aboriginal people (n=2,087). Compared to suicidal ideation only, a higher risk of repetition was observed for any self-harm method (aHR: 1.71; 95% CI: 1.37-2.12) amongst Aboriginal people and self-poisoning only (aHR: 1.45; 95% CI: 1.13-1.85) amongst non-Aboriginal people. Previous substance misuse was associated with a higher risk of repeat hospitalisation involving self-harm for Aboriginal (aHR: 1.7; 95% CI: 1.38-2.1) and non-Aboriginal (aHR: 1.6; 95% CI: 1.14-2.25) people. For non-Aboriginal people, several mental health diagnoses were associated with higher risks of repetition.

**Limitations:** The use of routinely collected administrative data limits analysis to only coded diagnoses and does not represent the full burden of self-harm and suicidal ideation in hospitals.

**Conclusion:** The similarities and differences in long-term risk of repeat hospitalisation involving self-harm between Aboriginal and non-Aboriginal people pose distinct challenges for clinical management and prevention. The results emphasise the importance of comprehensive psychosocial assessment to properly understand the interplay of individual and contextual influences and highlights the need to better understand the availability and effectiveness of culturally tailored clinical interventions and community-based solutions.

## Introduction

The Northern Territory (NT) of Australia has the highest rates of hospitalised self-harm [1] and deaths from suicide [2] of any jurisdiction in Australia. Self-harm is one of the strongest predictors of suicide [3], which makes it an important and urgent target for suicide prevention in the NT. Additionally, the repetition of self-harm is associated with further increasing the risk of suicide [4–6]. This has also been investigated in the NT in a cohort of hospital admissions involving self-harm and suicidal ideation, where the risk of suicide was found to increase by up to 30% for each repeat hospital admission involving self-harm [7]. Therefore, preventing repeat hospital admissions involving self-harm can contribute to reducing the risk of suicide in the NT.

Whilst there is evidence to inform measures that prevent the repetition of hospital-treated self-harm [8], its relevance to the unique context of the NT remains unclear. The NT has the highest proportion of Aboriginal and Torres Strait Islander (hereafter, respectfully referred to as Aboriginal) residents. Aboriginal people are the sovereign owners of and share collective and ancestral ties to the lands, waters, and skies that are intimately linked to their physical, psychological and spiritual wellbeing. They hold vital knowledge systems and expertise about health and wellbeing, but are grappling with the ongoing effects of colonisation contributing to health and social inequities and compromises in access to quality health care [9]. Aboriginal peoples continue to experience a higher risk of self-harm and suicide. A considered response is required which supports a self-determined cultural approach to address the complex systemic, social, historical, and political influences that are distinct from those affecting non-Aboriginal people in Australia [10]. However, only two recent cohort studies are known to have reported outcomes following hospitalisation involving self-harm, suicidal ideation, or both amongst Indigenous peoples from post-colonial countries similar to Australia. A cohort study from New Zealand found Māori were at higher risk compared to non-Māori of repeat presentation to ED involving self-harm, but this association was insignificant after adjusting for other demographic and clinical characteristics [11]. In a Canadian cohort study of children and adolescents hospitalised with self-harm the risk of repeat self-harm and suicide was higher amongst people who identified as American Indian or Alaska Native descent [12]. Whilst both studies suggest a greater risk of repeat self-harm exists amongst Indigenous peoples, they do not clarify the distinct risk factors that may need to be targeted for each of these population groups. Nor have these cohort studies considered suicidal ideation as an exposure, which has been identified as a potentially important expression of suicide risk amongst Aboriginal people in Australia [13,14].

Few longitudinal studies have highlighted risk factors for repeat hospitalisations involving self-harm amongst people initially hospitalised with suicidal ideation. In a study using Northern Ireland’s national self-harm registry surveillance data from episodes of hospitalisation involving both thoughts about and acts of self-harm, the risk of repeat hospital presentations involving self-harm was higher amongst individuals with an initial presentation involving suicidal ideation compared to self-harm [15]. Whilst the risk of repeat presentations involving self-harm was found to increase with the number of previous episodes of self-harm in a study from Spain, no differences in relative risk were observed between individuals presenting with self-harm and those presenting with suicidal ideation at the initial hospital presentation [16]. Further research is needed to bolster the mixed but limited evidence regarding the risk of self-harm following suicidal ideation in hospital settings.

In order to address these gaps in the evidence and inform efforts to reduce the population burden of self-harm in the NT a whole population retrospective cohort study was designed. Our aim with this study was to identify distinct risk factors amongst Aboriginal and non-Aboriginal people separately for repeat hospital admissions involving self-harm following an initial admission involving self-harm and/or suicidal ideation.

## Methods

### Study design and population

A retrospective cohort study was designed to include all NT residents with at least one hospital admission involving a diagnosis of self-harm (ICD-10-AM codes: X60-X84) and/or suicidal thoughts (ICD-10-AM codes: R45.81) between 1 July 2001 and 31 December 2013. The first hospital admission observed during the study period with a diagnosis of self-harm or suicidal ideation represented the index admission for longitudinal analysis. Individuals were excluded if, at the time of their index admission: no Indigenous status was recorded (n=2); their age was less than 10 years (n=10); or death occurred during admission (n=22). The follow-up period for each person in the study cohort was defined as the number of days from discharge at index admission until their next hospital admission involving self-harm or the first of any of the following censoring events: death, last recorded health service use in the NT, or end of follow-up on 31 December 2018.

### Data sources and collection

Individuals in the study cohort were linked to hospital records in the NT Inpatient Activity collection containing records of admissions to all NT public hospitals and to mortality records from the National Death Index (NDI) and the NT Deaths Registry. Details of these mortality data sources and how they were linked are described in more detail elsewhere [7].

The timestamps for last health service event in the NT were obtained from the Client Master Index (CMI). The CMI is a dataset established by the NT Department of Health to manage the Hospital Reference Number (HRN) used to uniquely identify individual users of any government-run health service, including public hospitals [17]. As part of the data management process, the CMI records the timestamp of the last service event by all individuals seen at any government-run health service. NT Department of Health staff deterministically linked each individual in the study cohort to their CMI records using the HRN obtained from hospital records collected for the study. Once each individual was linked, the timestamps for last health service event were extracted in 2020 and de-identified by replacing the HRN with encrypted project-specific keys that had been used for extracting the hospital and mortality records. These timestamps were then used for censoring individuals, as described above, for the purposes of analysis.

### Repeat hospital admissions involving self-harm

Repeat hospital admissions involving self-harm by any method were identified as records of hospital admissions during the follow-up period after the index admission containing any ICD-10-AM diagnosis code in the range of X60 to X84.

### Risk factors for repeat hospital admissions involving self-harm

#### Demographic characteristics

Sex, age, Indigenous status (coded as either Aboriginal or non-Aboriginal), and residence were obtained from the record of index admission. Administrative districts, established by the NT Department of Health for planning and organising health service delivery, was used to derive categories of residence. Residence was also categorised by region (Top End, for residents of the mostly tropical north of the NT from Katherine up to the capital city of Darwin; and Central Australia, for residents of the mostly arid areas in the south of the NT taking in the important service hub of Alice Springs) and remoteness (Urban, for residents of Alice Springs and Darwin Urban administrative districts; and Remote for residents in all other administrative districts) and a combination of region and remoteness (Top End Urban, Top End Remote, Central Australia Urban, and Central Australia Remote) to ensure the geographic distribution of the resident population can be adequately modelled with respect to how service delivery is organised.

#### Type of suicidal behaviour

Suicidal behaviour at the index admission was categorised as either suicidal ideation only or any type of self-harm. A more detailed categorisation distinguished between suicidal ideation and common methods of self-harm: self-poisoning (ICD-10-AM: X60-X69), self-cutting (ICD-10-AM: X78), hanging (ICD-10-AM: X70), and other types of self-harm (ICD-10-AM: X71-X77 and X79-X84). Where diagnosis codes of self-harm and suicidal ideation were co-present, admissions were coded into the relevant category of self-harm to ensure mutually exclusive categories. Where more than one type of method of self-harm was found, the admission was coded into the most lethal category of self-harm defined by national case fatality ratios [18].

#### Mental health conditions

Historical and current mental health conditions were derived from the diagnosis codes of prior and index hospital admission records, respectively. These codes were organised into mutually exclusive clinically relevant diagnosis groups that have been validated in other studies using administrative data from hospitals [19]. From the more refined diagnosis groups, aggregated categories of mental health conditions were also developed: severe mental disorders (comprising psychotic illnesses); common mental disorders (such as depression and anxiety); personality disorders; substance use disorders; other adult-onset disorders, and; other childhood onset-disorders. Other adult-and child-onset disorder categories were omitted from the final analysis due to small numbers that prevented meaningful analysis.

### Statistical analyses and procedures

A survival analysis approach was adopted to model the time to repeat hospital admissions involving self-harm associated with socio-demographic and clinical characteristics at index admission. Semi-parametric approaches were favoured to account for the distribution of survival times and right censoring observed. All analyses were stratified by Indigenous status to identify characteristics distinct to Aboriginal and non-Aboriginal populations. The Kaplan-Meier (KM) estimator was used to determine survival probability over time, which reflects the probability of not encountering a repeat hospital admission involving self-harm during follow-up. The absolute risk or probability of the outcome was determined by calculating 1-KM over the whole follow-up and at 1, 2, 5 and 10 years. Factors associated with a higher risk over time of repeat hospital admission involving self-harm were determined by hazard ratios (HR) estimated using multivariable Cox proportional hazards regression models. Adjusted Hazard Ratios (aHR) estimated for each covariate included in the final models were used to describe the relative risk over time of the outcome across different levels of the covariates. The final multivariable models were developed iteratively by including all candidate covariates from univariate analysis (see S1 Table and S2 Table in Supporting information) and removing one at a time where no strong evidence of an association could be observed.

All models retained adjustments by important population-level characteristics: sex, age at index admission, and residence. Residence was initially modelled using refined categories combining region and remoteness, but the small number of outcomes in some categories prevented the calculation of reliable estimates. Therefore, residence was modelled using region alone. Similarly, aggregated categories of mental health conditions were the preferred covariates used for developing multivariable models, with the broader categories being used to improve model performance or where all sub-categories were found to have similar estimated coefficients. The proportionality assumption of each covariate and the models overall were tested formally and visually using Schoenfeld residuals. The diagnostic value of the final multivariable regression models were measured using a pseudo R^2^ estimate of explained variation [20]. All analyses were undertaken using Stata 15.0 [21].

### Ethical approval

Ethical approval for this study was provided by the Human and Research Ethics Committees of the NT Department of Health and Menzies School of Health Research (Ref: 15-392) and the Australian Institute of Health and Welfare (Ref: 2013-3-31) for linkage to national mortality records used in this study for censoring.

## Results

During the study period, 4,391 individuals recorded an index hospital admission involving self-harm (n=2,956), suicidal ideation (n=1,292), or both (n=143). Just over half of the cohort (n=2,304; 52.5%) identified as Aboriginal, just under half (n=2,084; 47.5%) were female, and the median age at index admission was 30 years (IQR: 22-40). A more detailed summary of the demographic and clinical characteristics for Aboriginal and non-Aboriginal people is provided in the supporting information (see S1 Table and S2 Table).

During the follow-up period, 943 individuals were admitted on 1,985 occasions for self-harm (Aboriginal n=1,208; non-Aboriginal n=777). Of those with a repeat hospital admission involving self-harm, the number of episodes ranged from 1 to 28 (median: 1; IQR: 1-2). After accounting for censoring, 25.3% (95% CI: 23.8%-27.0%) of individuals in the study had experienced at least one repeat hospital admission involving self-harm (Aboriginal: 29.9%, 95% CI: 27.5%-32.4%; non-Aboriginal: 20.3%, 95% CI: 18.3%-22.4%). Figure 1 shows the survival probability (i.e., probability of not experiencing a repeat hospital admission involving self-harm) by Indigenous status illustrating a 40% higher risk of the outcome for Aboriginal compared to non-Aboriginal people over the course of the study (HR: 1.39; p < 0.001; 95% CI: 1.22-1.59). Table 1 provides a further summary of the distribution of absolute risk of repeat hospital admissions involving self-harm across demographic characteristics and type of suicidal behaviour at index admission for Aboriginal and non-Aboriginal people.

**Figure 1.**
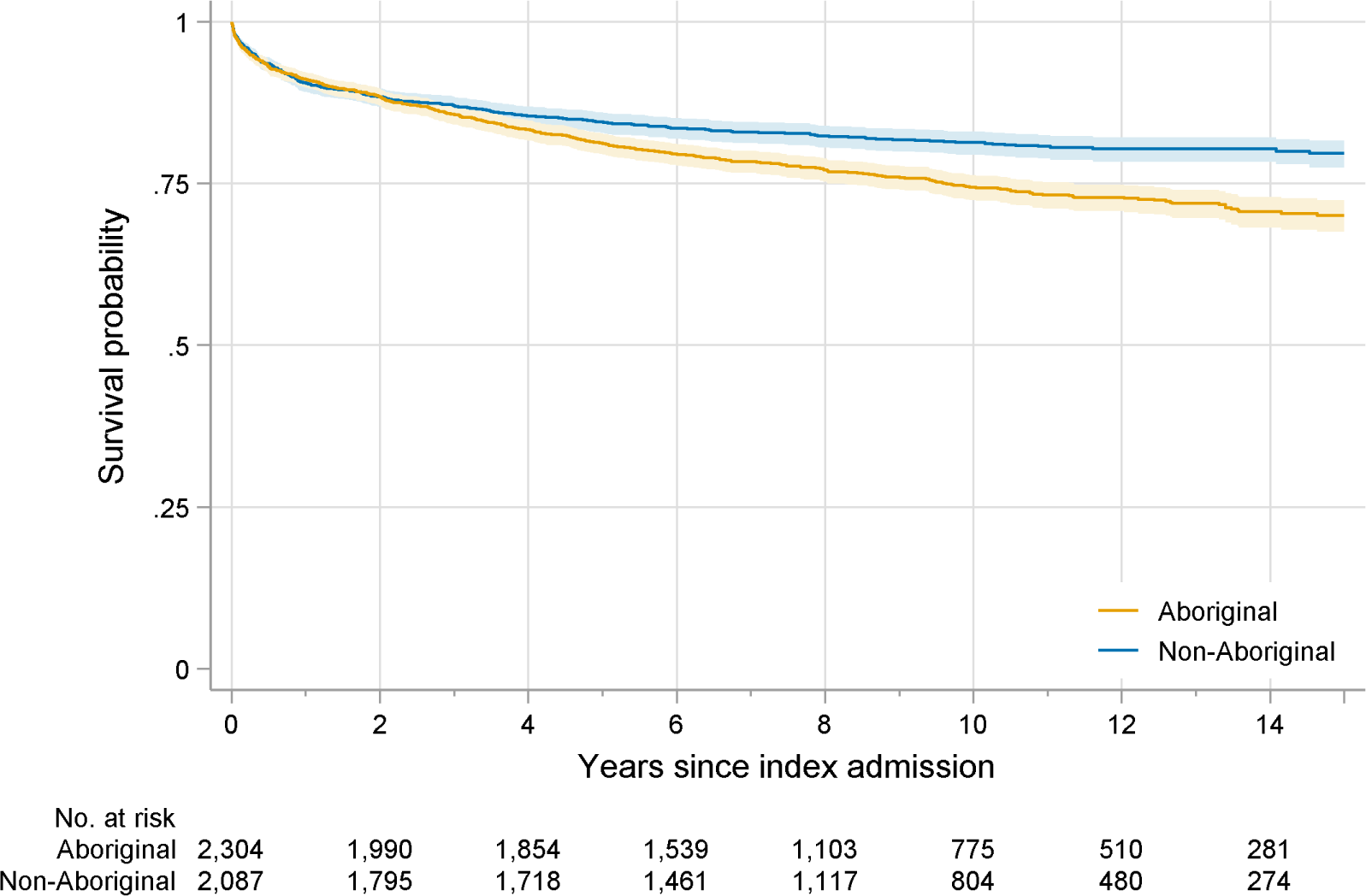
Probability of survival for Aboriginal and non-Aboriginal people following hospital admission involving self-harm and suicidal ideation, NT 2001-2013

**Table 1.** Descriptive summary and probability of repeat hospital admission for self-harm by Aboriginal and non-Aboriginal people with a hospital admission involving self-harm and suicidal ideation, NT 2001-2013.

|  | Individuals | Outcomes | Probability of outcome |  |  |  |
| --- | --- | --- | --- | --- | --- | --- |
|  | n | n | 1 year | 2 years | 5 years | Overall |
| **Aboriginal** |  |  |  |  |  |  |
| Total | 2,304 | 567 | 8.9% | 11.6% | 18.8% | 29.9% |
| *Sex* |  |  |  |  |  |  |
| Female | 1,131 | 286 | 9.6% | 12.3% | 19.2% | 32.1% |
| Male | 1,173 | 281 | 8.2% | 10.9% | 18.3% | 28.0% |
| *Age (years)* |  |  |  |  |  |  |
| 10-19 | 425 | 96 | 7.1% | 8.5% | 18.2% | 26.7% |
| 20-34 | 1,181 | 299 | 9.4% | 12.1% | 19.1% | 30.4% |
| 35-54 | 663 | 165 | 9.4% | 12.6% | 18.5% | 31.2% |
| 55+ | 35 | 7 | 5.8% | 11.9% | 18.0% | 22.1% |
| *Residence* |  |  |  |  |  |  |
| Central Aust. | 1,050 | 290 | 10.7% | 13.7% | 21.3% | 34.2% |
| Top End | 1,254 | 277 | 7.4% | 9.9% | 16.6% | 26.5% |
| *Type of suicidal behaviour* |  |  |  |  |  |  |
| Suicidal ideation | 628 | 100 | 5.0% | 6.1% | 12.2% | 21.5% |
| Self-harm | 1,676 | 467 | 10.4% | 13.6% | 21.2% | 32.6% |
| Self-poisoning | 440 | 113 | 9.4% | 13.3% | 21.2% | 28.8% |
| Self-cutting | 718 | 237 | 11.9% | 15.6% | 24.8% | 38.5% |
| Hanging | 379 | 81 | 7.7% | 10.1% | 15.5% | 26.2% |
| Other methods | 139 | 36 | 13.1% | 14.5% | 18.2% | 31.7% |
| **Non-Aboriginal** |  |  |  |  |  |  |
| Total | 2,087 | 376 | 9.4% | 11.6% | 15.5% | 20.3% |
| *Sex* |  |  |  |  |  |  |
| Female | 953 | 185 | 8.6% | 11.4% | 15.7% | 22.3% |
| Male | 1,134 | 191 | 10.1% | 11.8% | 15.2% | 18.3% |
| *Age (years)* |  |  |  |  |  |  |
| 10-19 | 346 | 65 | 9.6% | 13.0% | 16.2% | 19.9% |
| 20-34 | 784 | 132 | 10.0% | 11.1% | 14.4% | 19.0% |
| 35-54 | 763 | 156 | 9.6% | 12.5% | 17.3% | 23.4% |
| 55+ | 194 | 23 | 5.9% | 7.6% | 11.8% | 13.7% |
| *Residence* |  |  |  |  |  |  |
| Central Aust. | 508 | 103 | 11.1% | 13.3% | 17.9% | 22.2% |
| Top End | 1,579 | 273 | 8.9% | 11.1% | 14.7% | 19.7% |
| *Type of suicidal behaviour* |  |  |  |  |  |  |
| Suicidal ideation | 664 | 107 | 8.2% | 10.1% | 14.8% | 18.1% |
| Self-harm | 1,423 | 269 | 10.0% | 12.3% | 15.8% | 21.0% |
| Self-poisoning | 956 | 202 | 11.1% | 13.6% | 17.2% | 23.5% |
| Self-cutting | 262 | 49 | 10.0% | 13.1% | 16.2% | 20.2% |
| Hanging | 99 | 7 | 4.1% | 4.1% | 6.1% | 7.3% |
| Other methods | 106 | 11 | 4.8% | 6.7% | 10.7% | 10.7% |

In the multivariable modelling for both Aboriginal and non-Aboriginal people (Table 2), no strong evidence of substantial differences was observed in the relative risk of repeat hospital admission involving self-harm according to demographic characteristics, except for age in non-Aboriginal people. Compared to index admissions involving suicidal ideation alone, all methods of self-harm carried a higher risk of repeat hospital admission involving self-harm for Aboriginal people, whereas for non-Aboriginal people this was only the case for index admissions involving self-poisoning. Additionally, index admissions involving hanging by non-Aboriginal people had an approximately 2 times lower risk of repeat hospital admission involving self-harm compared to index admissions involving suicidal ideation. Diagnoses of substance use disorders, especially in relation to alcohol, were the most notable clinical risk factor associated with the outcome for both Aboriginal and non-Aboriginal people. For Aboriginal people, this association was found for diagnoses of alcohol use disorders in both index and previous hospital admissions, although the statistical support for the former was weak after adjusting for other factors. For non-Aboriginal people, diagnoses of alcohol use disorders at index admission and of substance use disorder in previous admissions were associated with repeat hospital admissions involving self-harm. Furthermore, diagnoses of severe mental disorders and personality disorders at index admission and diagnoses of common mental disorders in previous admissions were all independently associated with a higher risk of repetition for non-Aboriginal people. However, the pseudo R^2^ statistics estimated from the best fitting models (R^2^ : 0.097; R^2^ : 0.109) suggests that very little of the variance in the outcome was explained by the available data.

**Table 2.** Estimates from Cox regression models of repeat hospital admission involving self-harm by Aboriginal and non-Aboriginal people admitted to hospital with self-harm and suicidal ideation, NT 2001-2013.

|  |  | Aboriginal |  | Non-Aboriginal |  |
| --- | --- | --- | --- | --- | --- |
|  |  | HR | 95% CI | HR | 95% CI |
| **Demographic characteristics at index admission** |  |  |  |  |  |
| *Sex* |  |  |  |  |  |
| Female |  | 1 |  | 1 |  |
| Male |  | 0.87 | (0.73-1.03) | 0.95 | (0.76-1.17) |
| Age in years (median centred) |  | 0.99 | (0.98-1) | 0.99* | (0.98-0.99) |
| *Residence* |  |  |  |  |  |
| Top End |  | 1 |  | 1 |  |
| Central Australia |  | 1.13 | (0.94-1.34) | 1.13 | (0.9-1.42) |
| **Clinical characteristics at index admission** |  |  |  |  |  |
| *Type of suicidal behaviour* |  |  |  |  |  |
| Suicidal ideation |  | 1 |  | 1 |  |
| Self-harm |  | 1.71*** | (1.37-2.12) | 1.24 | (0.99-1.57) |
| Self-poisoning |  | 1.67*** | (1.27-2.2) | 1.45** | (1.13-1.85) |
| Self-cutting |  | 1.94*** | (1.53-2.47) | 1.14 | (0.81-1.6) |
| Hanging |  | 1.35* | (1.01-1.82) | 0.45* | (0.2-0.97) |
| Other methods |  | 1.62* | (1.1-2.38) | 0.71 | (0.38-1.33) |
| Severe mental disorders | No |  |  | 1 |  |
|  | Yes |  |  | 1.58** | (1.18-2.1) |
| Personality disorders | No |  |  | 1 |  |
|  | Yes |  |  | 1.83*** | (1.33-2.52) |
| Alcohol use disorders | No | 1 |  | 1 |  |
|  | Yes | 1.19 | (0.99-1.43) | 1.4** | (1.11-1.76) |
| **Clinical characteristics from previous admissions** |  |  |  |  |  |
| Common mental disorders | No |  |  | 1 |  |
|  | Yes |  |  | 1.58* | (1.1-2.26) |
| Substance use disorders | No |  |  | 1 |  |
|  | Yes |  |  | 1.6** | (1.14-2.25) |
| Alcohol use disorders | No | 1 |  |  |  |
|  | Yes | 1.7*** | (1.38-2.1) |  |  |

## Discussion

This is the first known cohort study to distinguish the risk of repeat episodes of hospital-treated self-harm by Indigenous status and one of a few that has compared outcomes between suicidal ideation and self-harm. The results point to several important implications for better understanding opportunities for preventing self-harm. Firstly, the lower absolute risk of repeat hospital admissions involving self-harm estimated in this study compared to others requires some clarification. The 1-year incidence of repeat hospital admissions involving self-harm in the NT we observed (8.9% and 9.4% amongst Aboriginal and non-Aboriginal people, respectively) is just over half of what was reported in a recent meta-analysis (17.0%) [22] and is the lowest reported in other hospital-based studies from Australia [23–25]. Study characteristics may explain this difference since studies that use hospital admissions data to define cohorts and outcome measures have tended to report a lower incidence of repeat episodes of self-harm [3]. This could be because the use of hospital admissions alone may be prone to underestimating the true incidence of hospital-treated self-harm due to the higher threshold of medical seriousness requiring admission [26,27]. Another factor could be the inclusion of suicidal ideation at inception, which is not common in other studies and which the results in this study show is associated with a lower risk of the outcome compared to self-harm at inception. However, of the methods associated with repetition from multivariable modelling, the estimated 1-year absolute risk of repetition for all self-harm amongst Aboriginal people (10.4%) and self-poisoning amongst non-Aboriginal people (11.1%) is still lower in this study compared to other studies. The relatively lower rate of repeat self-harm taken together with increasing trends in hospital admission involving suicidal ideation and self-harm in the NT observed during this study period [13] point to a potentially concerning increase in incident episodes that may suggest the need to also consider early intervention strategies to help prevent self-harm and subsequent suicide. However, it also possible that the results either point to potentially protective follow-up responses to hospital admissions involving suicidal ideation and self-harm or are an artefact of alternatives to hospital responses to repeat self-harm in community settings or in other parts of the healthcare system.

It should be noted that the absolute risk of repeat hospital admissions involving self-harm in the NT continued to increase over time compared to other studies, growing to around three quarters of the 5-year pooled incidence of repeat self-harm estimated by a meta-analysis of hospital-based studies (18.8% Aboriginal and 15.5% non-Aboriginal vs. 22.4% pooled estimated incidence) [3]. Whilst it has been suggested that the risk of repeat self-harm tends to plateau after 5 years [28], this study has shown continual long-term increases to almost 30% and just over 20% for Aboriginal and non-Aboriginal people, respectively. This suggests that the underlying risk of self-harm repetition observed in NT hospitals is likely the result of persistent or recurring long-term influences. In the absence of any association between enduring demographic characteristics that could explain these results, it is important that the broader context and longer-term influence of clinical characteristics are considered in drawing out implications for hospital management and further care in the community.

Substance misuse, especially involving alcohol, was especially prevalent and found to be associated with an increased risk of repeat hospital admissions involving self-harm. These results are consistent with existing evidence demonstrating a higher risk of a range of suicidal behaviours associated with alcohol use disorders [29]. Furthermore, the results appear to confirm both the proximal and distal influence of harmful alcohol consumption on the repetition of self-harm. That is, the association with previous admissions involving alcohol use disorders (for both Aboriginal and non-Aboriginal people) potentially points to the role of chronic misuse creating an underlying vulnerability to suicidal behaviour and the association with alcohol use disorders at index admission (non-Aboriginal people only) suggests that acute alcohol intoxication is a potential facilitator of suicidal behaviour and barrier to adopting alternative coping behaviours [30]. In light of the high rates of harmful alcohol consumption in the NT [31], the findings from this study reinforce the need for evidence-based population level policies to reduce harmful alcohol consumption that should contribute to reducing the risk of self-harm [32] and its repetition. Moreover, the very high burden of alcohol misuse on the hospital system in the NT [33] and the particularly high risk of self-harm associated with high levels of acute alcohol intoxication [34] suggests self-harm hospital admissions involving alcohol represent an important opportunity for referral to psychological interventions targeting misuse that are known to reduce the risk of subsequent self-harm in individuals [35]. Whilst further investigation is needed to determine the appropriateness and effectiveness of existing treatment services in the NT [36], it is even more important that mental health staff in the hospital are adequately supported to deal with the challenges posed by alcohol intoxication that often lead to poorer quality of care and non-assessment of self-harm [37].

The findings relating to mental health conditions diagnosed in non-Aboriginal people in the study point to important clinical risk factors that are known to increase vulnerability to self-harm [38] and help to clarify the potential opportunities for intervention. Psychological interventions, such as CBT, recommended for reducing vulnerability to self-harm repetition [39] should be considered as a therapeutic option as they have been shown to also effectively treat common mental disorders in self-harm, such as depression [40] and anxiety [41]. Furthermore, these treatments have also demonstrated effectiveness in reducing rates of self-harm repetition in the presence of personality disorder [42], especially borderline personality disorders, which this study suggests should also be a target for prevention in the NT. Since the NT has the highest rates of specialist mental health service use of any jurisdiction in Australia [43], further investigation into the quality and appropriateness of such care following hospitalisation for self-harm and suicidal ideation is warranted.

The association observed in non-Aboriginal people between repeat self-harm and severe mental disorders, comprising mostly schizophrenia and schizoaffective disorders, may require different approaches to clinical management depending on the stage and severity of psychosis or psychotic symptoms. The risk of self-harm is known to be greatest during early onset psychosis [44] and higher amongst those with other common risk factors for suicidal behaviour [45]. Furthermore, the existing evidence suggests that a higher risk of self-harm is associated with inadequate care and management in the community such as: poor adherence to medication [46], longer period of untreated psychosis [47,48], and higher frequency of psychiatric admissions [45]. These concerns may be particularly important for the NT where the use of depot antipsychotic medication for the treatment of psychotic illness has historically been the highest of any Australian jurisdiction [49]. The high use of depot medication may, among other things, reflect the application of clinical recommendations associated with widespread concerns with suicide risk in the NT clinical population [50]. For these reasons, comprehensive and integrated models of care, especially for people with early onset psychotic illnesses, are required that are known to assist with the management of significant mental health issues and reduce the risk of further suicidal behaviour [39].

The results from the stratified analysis point to distinct risks for repetition within Aboriginal and non-Aboriginal populations hospitalised for self-harm and suicidal ideation. The methods of self-harm associated with a higher risk of repetition differed by Indigenous status. Consistent with the evidence pointing towards higher risk of repetition associated with less fatal methods [51–53], self-poisoning was associated with a higher risk and hanging associated with a lower risk of repetition amongst non-Aboriginal people. For Aboriginal people, however, all methods of self-harm were associated with an increased risk of repetition. Since any form of self-harm repetition has been found to be associated with a higher risk of suicide [7], the findings from this study may point to overlaps between those at risk of repeat self-harm and suicide. Therefore, means restriction counselling to reduce the risk of repeat self-harm should be informed by comprehensive psychosocial assessment to ensure the effectiveness of such strategies are based on an understanding of the psychological and interpersonal contexts of intent and unmet needs. Younger age was associated with a higher risk of repetition for non-Aboriginal but not Aboriginal people, despite their younger age profile at index admission. Whilst the results for non-Aboriginal people are consistent with the evidence pointing to a higher frequency of repetition associated with the earlier onset of self-harm, especially in adolescence [54], issues of confounding or universally experienced influences may be at play for Aboriginal people. For example, it may be the case that clinically relevant associations, such as substance misuse and type of self-harm, may better explain age related differences in the results for Aboriginal people. Alternatively, it may be that Aboriginal people of all ages are universally affected by contextual influences not measured by this study, such as conditions of relative disadvantage, marginalisation and discrimination, cultural loss and intergenerational trauma [55]. However, the prevalence and role of alcohol misuse in this study more likely suggests the interplay of individual psychosocial stressors and underlying social determinants [56]. That is, the results from this study, taken together with Aboriginal perspectives, point to a combination of both individually experienced and collectively shared factors. Clinical management and aftercare for Aboriginal people should be informed by careful consideration of these influences, which requires culturally responsive assessment that balances both patient and cultural safety [57]. More importantly, investments in more culturally appropriate and safe services to effectively treat individuals in the community needs to be coupled with greater investments in self-determined and strengths-based prevention efforts targeting locally relevant issues and cultural supports [10]. Overall, this study suggests that the risk of repetition amongst non-Aboriginal people to be more closely associated with clinical populations for whom there is a need to ensure proper access to evidence-based treatments and effective services. But, for Aboriginal people, the higher risk of repeat hospital admission involving self-harm may be seen as the individual expression of adverse contextual influences requiring access to evidence-based individual treatments complemented by community-led cultural solutions.

### Strengths and limitations

This study makes a unique and important contribution to better understanding self-harm and suicidal ideation in the NT population, and amongst Aboriginal people in particular, with findings based on careful consideration of the distinctive social and cultural diversity of communities in the NT and availability of services. However, the uniqueness of the NT context means that caution must be used when generalising implications from these findings to other Australian settings or other Indigenous populations globally. Whilst the use of routinely collected administrative records for hospital admissions offers a high quality and reliable standard of data regarding demographic and clinical information, they do not represent all episodes of self-harm and suicidal ideation treated in the hospital setting. However, without reliable recording of self-harm and suicidal ideation in the emergency department, population-level insights to inform prevention can only make use of records for hospital admission [26]. Furthermore, the standardised coding of diagnoses does not include important interpersonal, social, and cultural influences that may or may not be recorded in hospital records and are especially important for understanding self-harm by Aboriginal people. This is likely one of the more important reasons for the limited explanatory value of clinical risk factors observed in this study. Further research that can access information typically gathered from psychosocial assessments and that privileges Aboriginal perspectives and knowledge are necessary to addressing these gaps.

## Conclusion

This study highlights important similarities and differences in long-term risk of repeat hospital admission involving self-harm for Aboriginal compared to non-Aboriginal people in the NT. The role of alcohol and substance misuse for both Aboriginal and non-Aboriginal people may reflect a complex intersection of individual vulnerability and specific psychosocial stressors that requires further investigation to better determine the role that hospitals may play in encouraging individual treatment and promoting prevention at a population level. For non-Aboriginal people the higher risk of repetition also appears to be associated with clinical populations, pointing to the need to better understand the availability of evidence-based interventions and the effectiveness of existing services in reducing risk and supporting recovery. There is also an urgent need to ensure that Aboriginal people can access culturally safe and responsive options for effectively managing and treating the immediate unmet needs of individuals after discharge. This should be complemented by greater investments in community-based interventions that promote culture, resilience, and self-determination that may better address the long-term underlying risk for self-harm. These findings highlight the challenges for prevention in the NT and reinforces the importance for comprehensive psychosocial assessment in hospitals to identify risk, needs, and strengths distinct to Aboriginal and non-Aboriginal people to better inform appropriate and effective options for care both within and beyond the hospital setting.

## Data Availability

The data used in the present study is not available outside of the investigators named on ethics approvals as per the conditions set by the owners/custodians of the data.

## Contributors

BL: Conceptualisation, methodology, formal analysis, data curation, writing – original draft, writing – review and editing, visualisation, project administration, funding acquisition; RB: Writing – review and editing; TH: Writing – review and editing; SS: Conceptualisation, methodology, writing – review and editing, funding acquisition; SG: Methodology, writing – review and editing; GR: Conceptualisation, methodology, writing – review and editing, funding acquisition.

## Acknowledgements

We would like to acknowledge the participants of this study who have had direct experience of suicide, including those who have attempted suicide and those bereaved by suicide. We would like to thank the advisory group to this study comprising Aboriginal and non-Aboriginal experts with clinical and lived experience for their generous insight and feedback. We thank the NT Department of Health for their support in obtaining and understanding the data for this study. The views expressed in this publication are those of the authors and not necessarily those of the funding body and partners to this study.

## Supporting information

**S1 Table. Descriptive summary and univariate analysis of repeat hospital admission involving self-harm by Aboriginal people with a hospital admission involving self-harm and suicidal ideation, NT 2001-2013**

**S2 Table. Descriptive summary and univariate analysis of repeat hospital admission involving self-harm by non-Aboriginal people with a hospital admission involving self-harm and suicidal ideation**

